# Revisiting VERTIGO and VERTIGO-CI: Identifying confidentiality breaches and introducing a statistically sound, efficient alternative

**DOI:** 10.1101/2025.05.30.25328653

**Authors:** Marie-Pier Domingue, Jean-François Ethier, Jean-Philippe Morissette, Simon Lévesque, Anita Burgun, Félix Camirand Lemyre

## Abstract

**Background:** Health Data Research Network Canada is tasked with facilitating large-scale health data research, such as statistical analyses that integrate, within a single model, data collected by different organizations, each holding distinct subsets of features corresponding to the same individuals, thereby forming a vertical data partition. To support logistic regression analyses in this setting, we assessed two recently proposed algorithms, VERTIGO and VERTIGO-CI, which enable parameter estimation and confidence interval computation, respectively, with respect to three aspects: the risk of re-identifying patient feature data, communication efficiency, and the extent to which model interpretability is preserved. This study has three main objectives: (1) highlighting confidentiality issues that arise with VERTIGO-CI, as well as those that may occur with VERTIGO when a data node holds only binary covariates; (2) reducing the number of required communication rounds; and (3) proposing an alternative (RidgeLog-V) to VERTIGO that excludes the intercept from the penalty term, which VERTIGO otherwise includes.

**Methods:** We inspected the quantities exchanged in the original algorithms and used linear algebra to identify reverse-engineering procedures that the coordinating center could employ to reconstruct raw data. We also analyzed the objective function of the optimization problem, leading to the proposal of an alternative formulation that requires only a single round of communication while allowing the intercept to be excluded from the penalty term.

**Results:** We showed that, when the VERTIGO-CI algorithm is executed, the coordinating center can reconstruct all individual-level data using simple vector-matrix operations. When the VERTIGO algorithm is executed and a data node has binary covariates only, the coordinating center may be able to recover individual data when parameter estimates are shared. We adapted the VERTIGO algorithm to reduce the number of communications and proposed a variant that excludes the intercept from the penalty term.

**Conclusions:** While the use of VERTIGO-CI, or of VERTIGO with binary covariates does not involve directly sharing raw data, confidentiality breaches may arise through reverse-engineering, illustrating that that the distributed nature of an algorithm does not inherently guarantee data privacy. This work also proposed a new algorithm (RidgeLog-V) that reduces operational costs and enhances model interpretability.

## 1 Background

As the health research and management community strives to improve care and ultimately health outcomes for citizens, the questions being asked require access to more diverse and extensive datasets. From genomic to environmental data, and from quantified-self information to clinical care data warehouses, the factors influencing health outcomes are often collected by various organizations and stored across different systems. Traditionally, the standard approach to analysis has been to centralize data by copying it to a single location. However, the growing variety, scale, and geographical distribution of the data needed often make this impractical. Legal, ethical, and social acceptability constraints frequently arise, preventing data from being shared or centralized. Health Data Research Network Canada (HDRN) is tasked with facilitating large-scale health data research, such as statistical analyses that integrate, within a single model, individual-level data collected by different organizations—for example, linking genomic data from participants in the Canadian Partnership for Tomorrow’s Health (CanPath) cohort [1] with their corresponding provincial medico-administrative records. This scenario illustrates a case of vertically partitioned data [2], where different institutions hold distinct subsets of features corresponding to the same individuals. In such settings, the intended analyses often involve estimating parameters of statistical models and their associated confidence intervals (CIs). To support these objectives, we explored methods designed to accommodate such data-sharing constraints across institutions in the context of vertically partitioned data, with a focus on logistic regression—widely used in health research for modeling binary outcomes.

The field of distributed analysis offers relevant approaches in which analytical results are produced solely through the exchange of summary statistics. However, the absence of direct raw data sharing does not inherently guarantee confidentiality, as participating entities may still infer sensitive information by reverse-engineering the exchanged quantities, a risk that has been illustrated in the literature [3], in a context however different from the one considered in this paper.

In supporting vertically distributed logistic regressions, the ideal method does not require any raw data to leave the environments of the original data stewards (data nodes); does provide formal confidentiality guarantees, including protection against reverse-engineering; offers documented use in clinical research settings; is communication-efficient, as most data partners require human review of any information leaving a secure environment; and yields results that are comparable to those from classical pooled, maximum likelihood-based analyses (see e.g. [4]), which is important to reassure colleagues that the method is a valid alternative to the centralized approaches they have relied on until now. However, our review of the literature did not identify any method that satisfies all these criteria.

To our knowledge, only one method [5, 6] has been proposed that addresses the maximum likelihood-based logistic regression inference problem considered here, while fully avoiding the need for raw data to leave its original environment (while CIs are not specifically covered, it is straightforward to see that they can be obtained by a minor modification of the procedure described). However, as noted by the original authors and in a recent review [7], it requires a high number of communication rounds between parties, and concerns about data leakage persist—issues that may be inherently difficult to resolve. Taken together, these limitations rendered the method unsuitable for our intended applications.

The next step was to identify a method with minimal data-sharing requirements. The VERTIGO algorithm (JAMIA 2016) [8] and its extension developed by the same group, VERTIGO-CI (AMIA 2021) [9], have recently been proposed for the analysis of vertically partitioned data. They have since been applied in distributed health data research [10], including a study published in *Nature Communications* [11]. To implement the method, the vector of binary responses must be accessible to all involved parties, including the coordinating center (CC), which is responsible for aggregating the quantities shared by parties contributing with local data (called *nodes*). While this requires sharing the outcome outside the originating site, we hypothesized that it might be helpful to support some projects at HDRN if the other criteria were met, given the authors’ assertion that the VERTIGO algorithm “harmonize[s] information in a privacy-preserving manner”[9].

In order to ensure due diligence with respect to HDRN data partners, we conducted an assessment of VERTIGO and VERTIGO-CI regarding the risk of re-identifying patient feature data, the communication efficiency, and the extent to which they preserve the integrity of model interpretation. Four areas of concern were identified:

- **Confidentiality:** The use of VERTIGO-CI enables reverse engineering at the CC level, meaning that the quantities exchanged by local data nodes allow the CC to reconstruct individual feature data using simple vector-matrix operations;
- **Confidentiality:** The use of VERTIGO (estimation only, without CIs) may allow the CC to reconstruct individual feature data when a data includes holds binary features and discloses associated parameter estimates;
- **Communication:** The use of VERTIGO (estimation only, without CIs) involves non-essential communication rounds between local data nodes and the CC;
- **Comparability:** The use of VERTIGO (estimation only, without CIs) incorporates the intercept in the penalty term. However, in most applications of penalized logistic regression, the intercept is excluded from the penalty [12]. As a result, VERTIGO estimates are not directly comparable to those obtained from widely adopted algorithms such as the one proposed in [13], leading to inconsistencies in interpretation.

In light of these observations, the primary aim of this article is to guide the future use of this method. Specifically, our objectives are to: (1a) highlight the confidentiality issues associated with VERTIGO-CI and (1b) the privacy breaches that may arise from estimates disclosed when a node includes binary features under VERTIGO, (2) propose an optimized way to perform the operations on shared quantities to reduce the number of communication rounds, and (3) propose an alternative approach to VERTIGO that excludes the intercept from the penalty.

While the method discussed below is supported by mathematical arguments, we also illustrate the different aims with R code [14] available at https://github.com/OpenLHS/Revisiting VERTIGO using a publicly available dataset that was previously analyzed in the original VERTIGO-CI paper. For issues 1a and 1b, we numerically demonstrate how vector-matrix operations can be used by the CC to reconstruct data from other nodes using shared quantities.

The implementation of our alternative approach—named RidgeLog-V (Ridge Logistic Regression from Vertically Partitioned Data)—which is more communication-efficient and excludes the intercept from the penalty, is also available in the same repository. We illustrate that RidgeLog-V produces results that are coherent—yielding near-identical estimates to those obtained using glmnet [13]—while differing from those produced by the VERTIGO algorithm.

## 2 Methods

### 2.1 General methodological framework and mathematical foundations

The work presented here is expressed using a mathematical notation framework aligned with the one proposed in our previous work on horizontally distributed analyses [15]. The general methodological framework, including mathematical notations, is described below.

#### 2.1.1 Mathematical notations and definitions

In the following, non-italic uppercase letters represent random variables, while non-italic bold uppercase letters denote random vectors. Vector-valued quantities and matrices are represented by lowercase italic bold letters and uppercase italic bold letters, respectively. The *j*^th^ element of a vector ***a*** ∈ ℝ^*p*^ is denoted by *a*_*j*_. For any vector ***a*** ∈ ℝ^*p*^, diag(***a***) represents the *p* × *p* diagonal matrix with the entries of ***a*** on the diagonal. In the case of iterative computations over ***a***, the step count *s* appears as an index in parentheses (e.g., ***a***_(*s*)_). The gradient of a function *f* (***θ***) with parameter ***θ*** evaluated at ***θ*** = ***a*** is denoted as the column vector ∇_***θ***_*f* (***a***) := ∇_***θ***_*f* (***θ***)|_***θ***=***a***_. Notation glossaries are provided in Additional file 1-A.

We use the following formulation of the binary logistic regression model with *p* covariates. Let Y ∈ {−1, 1} be a random variable representing the binary response, and let the corresponding random vector of covariates be denoted by **X** ∈ ℝ^*p*^. The conditional distribution of Y | **X** = ***x*** is given, for *y* ∈ {−1, 1}, by

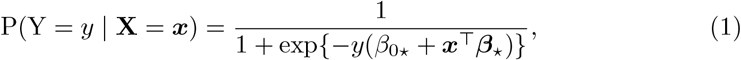

where *β*_0*⋆*_ ∈ ℝ and ***β***_*⋆*_ = [*β*_*⋆*1_, …, *β*_*⋆p*_]^⊤^ ∈ ℝ^*p*^ are respectively the true unknown intercept and the true unknown covariate parameters. The data involved in the analysis consists in a sample of *n* realizations of the random vector (**X**, Y), denoted by *𝒟* := {(***x***_1_, *y*_1_), …, (***x***_*n*_, *y*_*n*_)}, with ***x***_*i*_ = [*x*_*i*1_, …, *x*_*ip*_]^⊤^.

In the definition of the logistic model above, we adopt a different notation convention for the intercept parameter compared to the original VERTIGO and VERTIGO-CI articles. Specifically, we separate the intercept parameter from the covariate-associated parameters. In the original papers, a value of 1 is included in the covariate vector **x**, and the intercept parameter is treated as part of the covariate-associated parameters.

We represent the covariate data of the *n* individuals in matrix form and the response data in a vector form such that

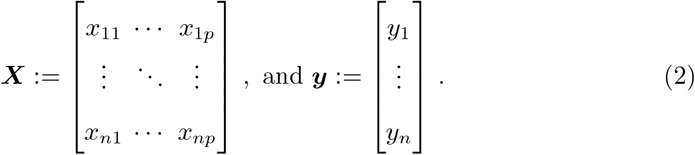

Our derivations involve the Gram matrix ***𝒦*** defined as follows, where **1**_*n*_ ∈ ℝ^*n*^ denotes the vector with all entries equal to one:

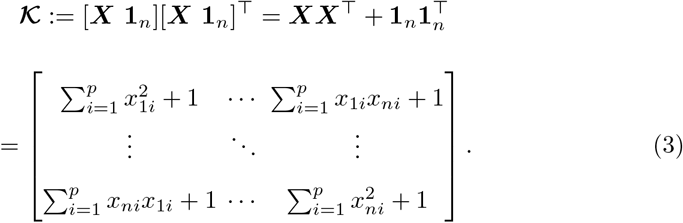

The (*i, j*) entry of **𝒦** for *i, j* ∈ {1, …, *n*} will be denoted *𝒦*_*ij*_. It can be readily deduced from (3) that the following holds for all *i, j* ∈ {1, …, *n*} :

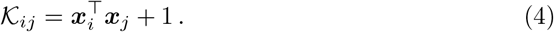

#### 2.1.2 Vertically partitioned data setting and properties of the Gram matrix

In the case of vertically partitioned data, where covariate data are distributed across *K* locations (referred to as nodes), the matrix ***X*** is partitioned into *K* submatrices denoted as ***X***^(1)^, …, ***X***^(*K*)^. Each node *k* ∈ {1, …, *K*} holds a matrix 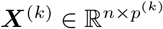 containing *p*^(*k*)^ covariates, ensuring that 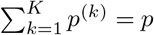. We assume that the columns of ***X*** are arranged such that the first *p*^(1)^ columns correspond to the covariates stored at node 1, followed by the next *p*^(2)^ columns corresponding to those stored at node 2, and so forth. Under this ordering, ***X*** can be expressed in a block matrix notation as

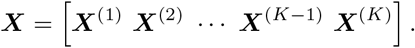

For *k* ∈ {1, …, *K*}, the *i*^th^ row of ***X***^(*k*)^, which we will denote by 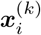, represents the *p*^(*k*)^ covariate values of individual *i* stored at node *k*. Using (4), it follows that

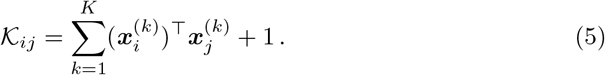

In VERTIGO and VERTIGO-CI, each node *k* ∈ {1, …, *K*} is assumed to have access to the response vector ***y*** in addition to the covariate data ***X***^(*k*)^. It is also assumed that the datasets are aligned across all partitions ***X***^(1)^, …, ***X***^(*K*)^ and that this alignment corresponds to the structure of ***y***, i.e., Patient 1 appears in row 1 of all partitions and in the first entry of ***y***, and so forth. We will maintain these assumptions throughout.

#### 2.1.3 Penalized logistic regression

VERTIGO and VERTIGO-CI rely on a penalized maximum likelihood estimation procedure, specifically the ridge logistic regression [16], to estimate the unknown coefficients *β*_0*⋆*_ and ***β***_*⋆*_ in (1). A strictly positive parameter *λ* is used to control the strength of the penalty. In these algorithms, the penalty is applied to all model coefficients, including the intercept. The corresponding optimization problem is given by

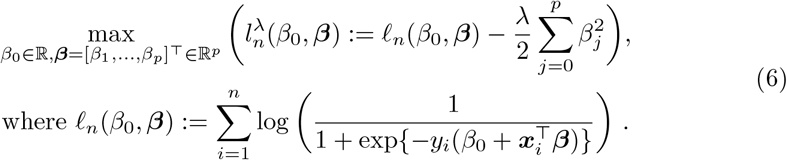

For *λ >* 0, the optimization problem in (6) always has a unique solution. Let 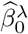 and 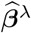 denote this solution.

As mentioned in the introduction, in most applications, the intercept is excluded from the penalty term [13]. When this is the case, the following alternative penalized maximum log-likelihood optimization problem is used to derive the model parameter estimates:

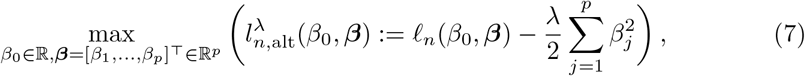

where *ℓ*_*n*_(*β*_0_, ***β***) is as in (6). For *λ >* 0, when there is at least one *y*_*i*_ equal to 1 and one *y*_*i*_ equal to 0, the optimization problem in (7) always has a unique solution (see Additional file 1-D). Let 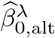 and 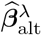 denote this solution.

### 2.2 Overview of VERTIGO

The VERTIGO algorithm leverages an application of dual optimization techniques, which shows that, under our notations, the unique solution 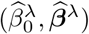 to the problem in (6) can be computed as

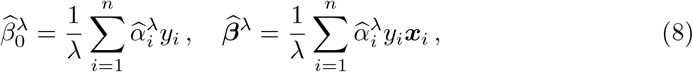

where 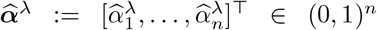 is the unique solution to the following minimization problem

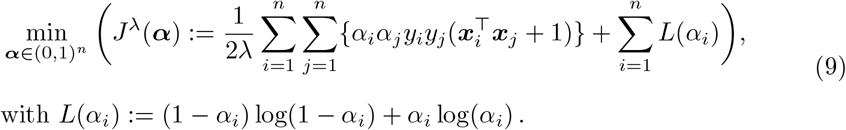

The minimization problem in (9) is called the dual problem to the maximization problem in (6), where the vector ***α*** is the dual parameter.

To compute 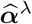 in the vertically partitioned setting described above, the VERTIGO algorithm employs an iterative Newton method. At each iteration, the gradient and Hessian of *J*^*λ*^(***α***) with respect to ***α*** can be computed exactly at the CC by requiring each node to send intermediate numerical outputs. Specifically, for *k* ∈ {1, …, *K*}, letting ***e***^(*k*)^(***α***) denote a vector whose *j*^th^ entry is given by

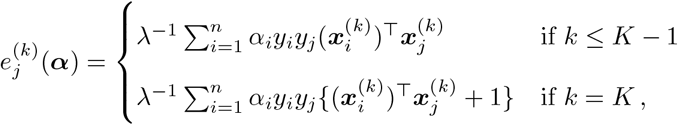

the gradient of *J*^*λ*^(***α***) in (9) is expressed as

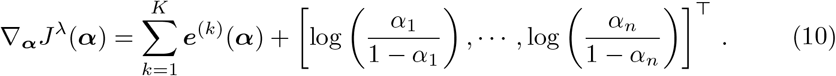

The distinct definitions for nodes *k* ∈ {1, …, *K* − 1} and node *K* reflect the fact that, in VERTIGO, a column of ones is appended to the data matrix ***X***^(*K*)^ at node *K* to treat the intercept as part of the covariate-associated parameters.

The Hessian of *J*^*λ*^(***α***) is expressed as

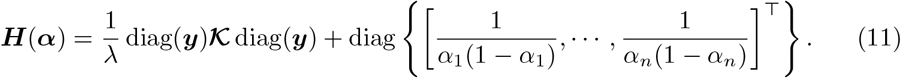

At the start of the VERTIGO algorithm, each node *k* shares the local matrix **𝒦**^(*k*)^ with the CC, where

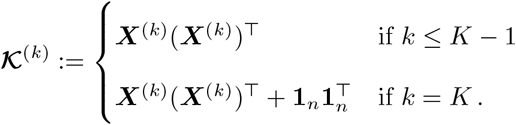

This allows the CC to construct **𝒦** because 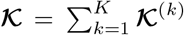(from (5)). Starting with ***α***_(0)_ shared across all nodes, at each iteration *s* ≥ 0 until convergence, each node sends its locally updated quantity ***e***^(*k*)^(***α***_(*s*)_) to the CC. The CC then updates ***α***_(*s*)_ using the Newton-Raphson algorithm such that ***α***_(*s*+1)_ = ***α***_(*s*)_ − ***H***^*−*1^(***α***_(*s*)_)∇_***α***_*J*^*λ*^(***α***_(*s*)_), and subsequently sends the updated dual parameters to all nodes.

Once the optimal dual parameter 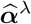 has been obtained and shared with all nodes, each node *k* computes the components of 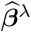 corresponding to the covariates stored at their node using the equation 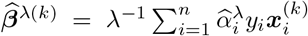 where this relationship follows from (8). The coefficient 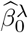 is computed at node *K* using the formula 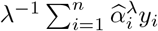

#### 2.2.1 Overview of VERTIGO-CI

The VERTIGO-CI algorithm [9] is an extension of the algorithm VERTIGO that aims at estimating the variance-covariance matrix of the estimator 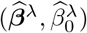 using {[***X* 1**_*n*_]^⊤^***V***^*λ*^[***X* 1**_*n*_]}^*−*1^, where ***V***^*λ*^ denotes a diagonal matrix whose entry (*i, i*) is given by

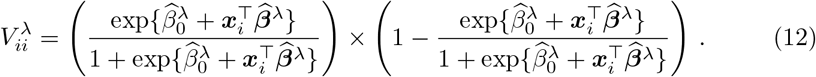

To achieve this, the VERTIGO-CI algorithm (provided in Additional file 1-B, see Algorithm 4 therein) begins by executing the VERTIGO algorithm to obtain 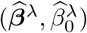 with a simplified Hessian matrix. Once the parameter estimates obtained, the nodes compute the matrix ***V***^*λ*^ using node-to-node communications (step 8 Algorithm 4 in Additional file 1-B). Every node can then compute and send (***X***^(*k*)^)^⊤^(***V***^*λ*^)^1*/*2^ to the CC ([***X***^(*k*)^ **1**_*n*_]^⊤^(***V***^*λ*^)^1*/*2^ for *k* = *K*). Finally, the CC builds the matrix [***X* 1**_*n*_]^⊤^***V***^*λ*^[***X* 1**_*n*_] by block.

The quantities shared between the data nodes and the CC throughout the algorithms VERTIGO and VERTIGO-CI are summarized in Table 1.

**Table 1.** Overview of the information shared in VERTIGO and VERTIGO-CI.

| Step | From nodes to CC | From CC to nodes |
| --- | --- | --- |
| Initialization<sup>1</sup> | $\mathcal{K}^{(k)}$ | - |
| Every iteration $s$ | $\mathbf{e}^{(k)}(\alpha_{(s)})$ | $\alpha_{(s)}$ |
| Estimation results | $\widehat{\beta}^{\lambda(k)}$ | - |
| CIs (VERTIGO-CI) | $(\mathbf{X}^{(k)})^\top (\mathbf{V}^\lambda)^{1/2}$ | - |

<sup>1</sup> It is assumed that all parties (nodes and CC) have access to $\mathbf{y}$ and $\lambda$ .

### 2.3 Methodology pertaining to objective 1

To highlight the confidentiality issues of VERTIGO-CI, using the list of available quantities at the CC in Table 1, we demonstrate how one can construct an invertible system of equations that, through linear algebra manipulations, enables the recovery of each node’s data matrix ***X***^(*k*)^.

To highlight the privacy breaches that may arise when a node includes binary variables and shares the associated parameter estimates under VERTIGO, we present a case in which the CC is able to reverse-engineer individual-level data for ***X***^(*k*)^ involving solely binary features. This is achieved using the local Gram matrix **𝒦**^(*k*^) and the estimated coefficients 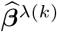 shared from node *k*. The case also relies on the following property of binary matrices, which can be established using basic tools from linear algebra:

Let ***A*** denote an *n*×*p* matrix with binary (0-1) entries. Consider ***a*** ∈ℝ^*n*^ and ***d*** ∈ ℝ^*p*^ such that ***a*** = ***Ad***, and let U(***d***) := {*z* ∈ℝ : *z* = ***b***^⊤^***d, b*** ∈ {0, 1}^*p*^}. If *𝒰* (***d***) has cardinality 2^*p*^, then any binary matrix ***A***^*′*^ satisfying ***a*** = ***A***^*′*^***d*** must be equal to ***A***.

### 2.4 Methodology pertaining to objective 2

Our optimization of the VERTIGO algorithm’s execution relies on the observation that, as the term 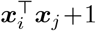in the definition of *J*^*λ*^(***α***) in (9) is equal to *𝒦*_*ij*_ (see equation (4)), *J*^*λ*^(***α***) can be expressed as

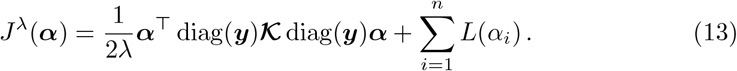

Based on this observation (details in Additional file 1-C), we designed an algorithm that requires only a single communication from the nodes to the CC, in which the nodes transmit the same quantities they send in Step 1 of the VERTIGO algorithm.

### 2.5 Methodology pertaining to objective 3

The design of RidgeLog-V, our proposed alternative approach to VERTIGO which excludes the intercept from the penalty, leverages dual optimization techniques very similar to those used in VERTIGO to derive the expression of *J*^*λ*^(***α***). These techniques show that, if there exists at least one *i* such that *y*_*i*_ = 1, at least one *j* such that *y*_*j*_ = −1 and *λ >* 0, the solution 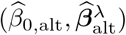 of the optimization problem (7) is unique and satisfies:

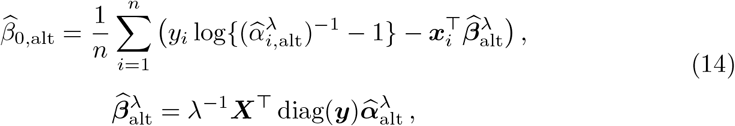

where 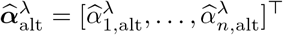 is the unique solution to the following minimization problem:

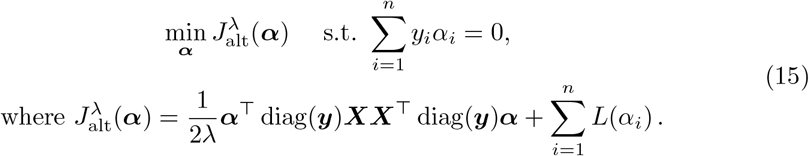

The proof of the latter result is available in Additional file 1-D.

To show how 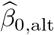 can be computed in our vertically partitioned data setting, we will also use the fact that as (14) implies 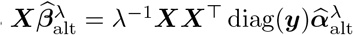, it holds that

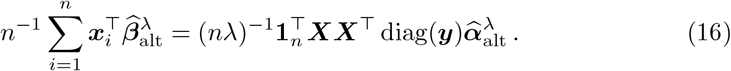

## 3 Results

### 3.1 Results pertaining to objective 1: Confidentiality

We first show that once the VERTIGO-CI algorithm has been executed, the quantities available at the CC allow it to reverse-engineer the feature data of all individuals. We then illustrate the privacy risks that can arise when binary features are used under VERTIGO and their associated parameter estimates are shared (even without using the CI extension).

#### 3.1.1 VERTIGO-CI

Once VERTIGO-CI has been executed, the following quantities are available at the CC. The corresponding steps (indicated in parentheses) refer to those shown in the VERTIGO-CI algorithm reproduced in Additional file 1-(B) (see Algorithm 4):

- The response vector ***y*** and the parameter *λ* (as part of the input of the algorithm);
- The local matrices **𝒦**^(1)^, …, **𝒦**^(*K*)^ (Step 1);
- 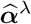 (Step 4);
- (***X***^(1)^)^⊤^(***V***^*λ*^)^1*/*2^, …, (***X***^(*K*)^)^⊤^(***V***^*λ*^)^1*/*2^ (Steps 9 & 10).

We next show how the CC can reconstruct the individual-level data held by each node using these quantities. From the relationships in (8), it holds, for all 1 ≤ *j* ≤ *n*, that

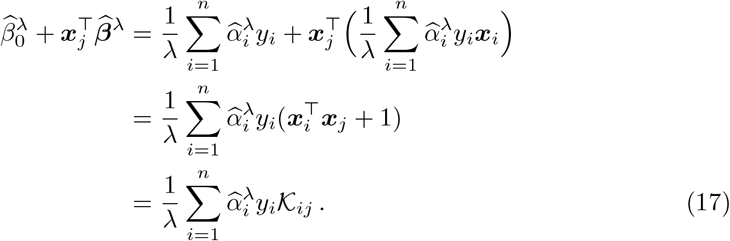

To obtain the last equality, we used the relationship at (4). Since the CC can calculate each entry of the Gram matrix 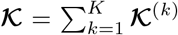 (it does so at Step 2), and as it has access to 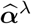, ***y*** and *λ*, the CC is able to compute 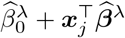 for each *j* ∈ {1, …, *n*} through the formula proved at (17).

Because it can compute 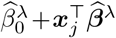 for each *j* ∈ {1, …, *n*}, the CC is therefore able to compute each entry of the diagonal matrix ***V***^*λ*^ defined in (12) through the formula

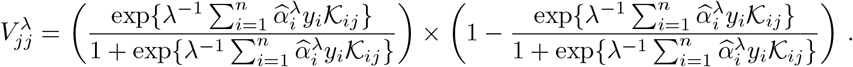

The last formula implies that the CC is also able to compute the inverse of the matrix (***V***^*λ*^)^1*/*2^. Indeed, since ***V***^*λ*^ is diagonal with strictly positive entries, (***V***^*λ*^)^*−*1*/*2^ is also a diagonal matrix whose *j*th diagonal entry can be calculated by the CC using 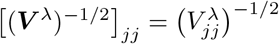.

Recalling that the CC has access to (***X***^(*k*)^)^⊤^(***V***^*λ*^)^1*/*2^ for each *k* ∈ {1, …, *K*}, and given that we have just shown how the CC can construct the matrix (***V***^*λ*^)^*−*1*/*2^ using quantities available to it after the execution of the VERTIGO-CI algorithm, we conclude that the CC can reverse-engineer each data matrix ***X***^(*k*)^ via the relationship

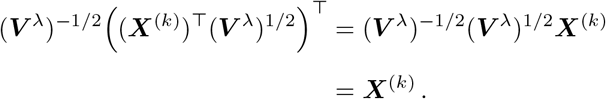

This is not the only way for the CC to proceed with reverse-engineering as another path can be used to construct the matrix ***V***^*λ*^ at the CC (see Additional file 1-E for details).

The reverse-engineering process is numerically illustrated using R code available at https://github.com/OpenLHS/RevisitingVERTIGO.

#### 3.1.2 Binary features in VERTIGO

We now elaborate the case in which the CC is able to reverse-engineer an individual patient data matrix ***X***^(*k*)^ comprising binary (0-1) features. We demonstrate it for a case with all binary predictors but mix cases (binary and continuous) warrant additional investigations before confirming or infirming their confidentiality properties.

For simplicity, to avoid dealing with the intercept in our example, we choose *k* ≠ *K*. We note that the relationship in (8) implies

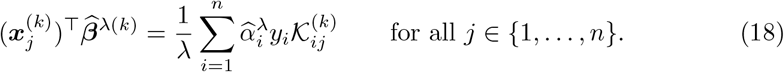

Since the CC has access to the matrix **𝒦**^(*k*)^ and the response vector ***y*** as part of the VERTIGO algorithm’s initialization, and also has access to 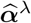, which is computed at the CC, it can compute (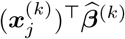 for all *j* ∈ {1, …, *n*} using formula (18). Therefore, when the parameter estimates 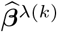 are made available to the CC, the only quantity unknown to the CC in (18) are the entries of 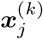. Using a vector-matrix notation, the CC can attempt to reverse-engineer the data matrix ***X***^(*k*)^ by identifying a binary matrix ***A*** such that 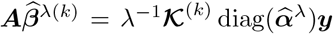. When the features of data node *k* are all binary, even if the number of unknowns exceeds the number of equations, if the cardinality of the set 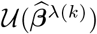 is equal to 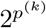, then any binary matrix satisfying 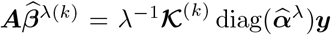 must be equal to ***X***^(*k*)^. In this case, to identify line *j* of ***X***^(*k*)^, the CC has to search over all binary vectors 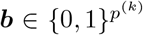 and identify the unique one such that 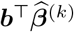 equals the *j* component of 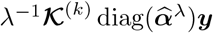.

To illustrate this, consider the case *p*^(*k*)^ = 3, and suppose that, upon running the algorithm, the vector 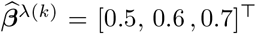 is obtained. In this case, it is straightforward to verify that the cardinality of the set 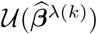 is 8 = 2^3^, and that the only possible values for the entries of 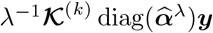 are

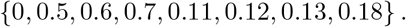

Therefore, if, for example, the *j*^th^ component of the vector 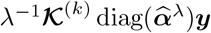 is equal to 0.12, the only possible value for the *j*^th^ row of ***X***^(*k*)^ is [1, 0, 1]^⊤^. The CC can then repeat this process to reconstruct the entire matrix ***X***^(*k*)^.

These confidentiality issues are numerically illustrated using R code available at https://github.com/OpenLHS/RevisitingVERTIGO.

### 3.2 Results pertaining to objective 2: Communication

The dual objective function used in VERTIGO, and its associated gradient and Hessian with respect to ***α***, only depends on the data through **𝒦** and ***y***. It follows that optimization procedure used to calculating **𝒦** can be entirely executed at the CC with the knowledge of **𝒦** and ***y***. To illustrate, we develop the objective function, the gradient and the Hessian in a way that highlights that only ***y*** and ***X***^(*k*)^(***X***^(*k*)^)^⊤^ for all *k* ∈ {1, …, *K*} are needed for the CC to complete the estimation procedure.

First, since 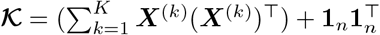 (see (5)), it directly follows that

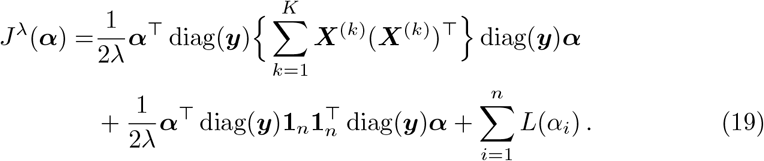

Consequently, if each node *k* sends the matrix ***X***^(*k*)^(***X***^(*k*)^)^⊤^ to the CC, and assuming the CC has access to the response vector ***y***, the CC can fully reconstruct the objective function of the optimization problem in (9). As will be shown, the CC is also able to solve this problem and compute 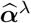 using Newton iterations without requiring any additional communication with the nodes.

From (10), since

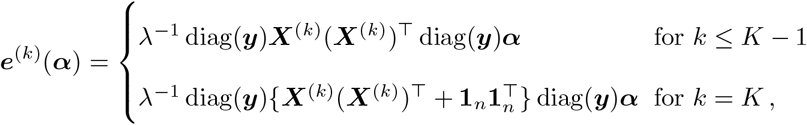

the gradient of *J*^*λ*^(***α***) can be expressed as

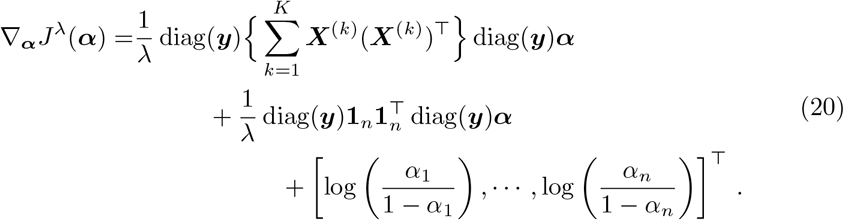

This expression for the gradient can also be derived directly from (19) using standard properties of vector calculus.

Moreover, from (11), the Hessian matrix of *J*^*λ*^(***α***) expresses as

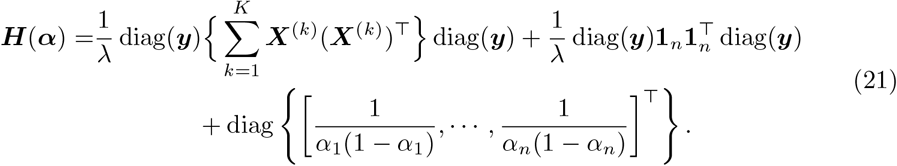

The expressions for ∇_***α***_*J*^*λ*^(***α***) and ***H***(***α***) the matrices ***X***^(*k*)^(***X***^(*k*)^)^⊤^, it is able to compute the Newton update without any additional communication with the nodes.

The version of the method with optimized communication is presented in Algorithm 1. The equivalence between the two algorithms follows from the fact that their Newton updates are mathematically identical. The information exchanged during the communication steps in the optimized version of the algorithm is summarized in Table 2.

**Table 2.**
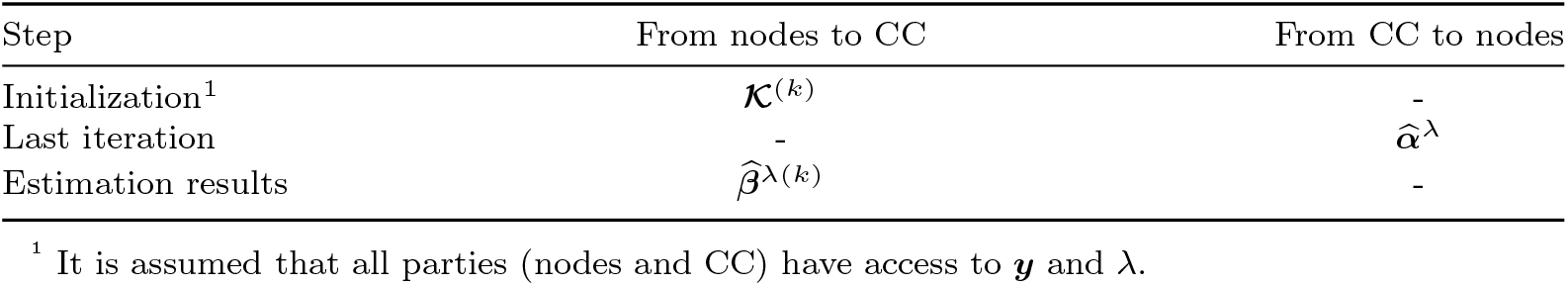
Overview of the information shared in the communication optimized algorithm.

| Step | From nodes to CC | From CC to nodes |
| --- | --- | --- |
| Initialization<sup>1</sup> | $\mathcal{K}^{(k)}$ | - |
| Last iteration | - | $\hat{\boldsymbol{\alpha}}^\lambda$ |
| Estimation results | $\hat{\boldsymbol{\beta}}^{\lambda(k)}$ | - |

<sup>1</sup> It is assumed that all parties (nodes and CC) have access to $\mathbf{y}$ and $\lambda$ .

#### Algorithm 1

Optimized communications

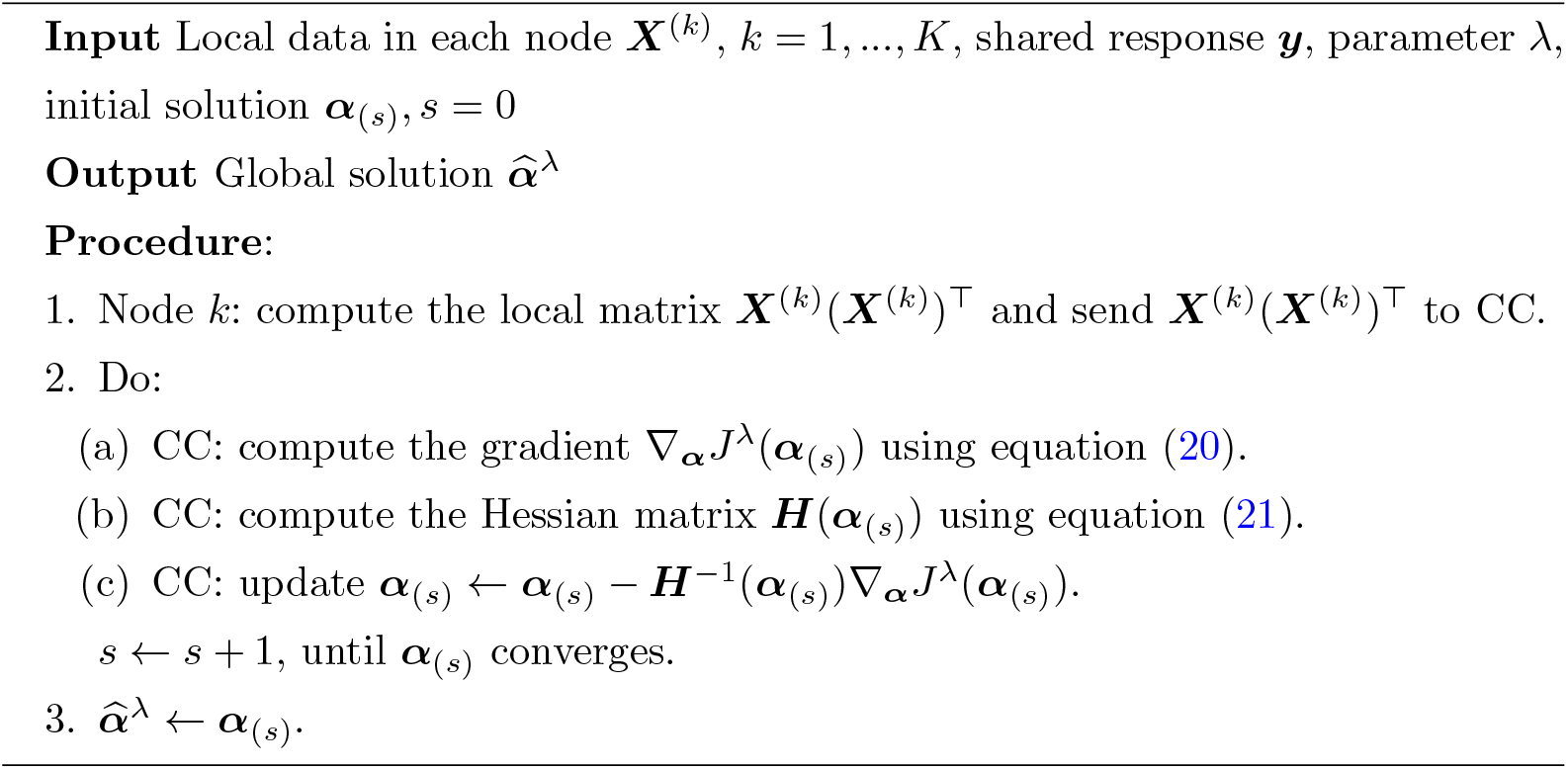

### 3.3 Results pertaining to objective 3: Comparability

The alternative approach proposed, which excludes the intercept from the penalty, is outlined in Algorithm 2. Once each node has sent the matrix ***X***^(*k*)^(***X***^(*k*)^)^⊤^ to the CC, the constrained dual problem in (15) is solved at the CC level using constrained optimization techniques that require no further communication or information exchange between the nodes and the CC. Multiple constrained convex optimization algorithms exist and some are implemented in software packages, such as the CVXR package in R [17]. We refer to [18] for further information regarding constrained optimization. After computing 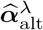, the CC broadcasts 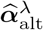 to all nodes. From this, to obtain the covariate-associated coefficients, the nodes use equation (14) with their locally stored data such that

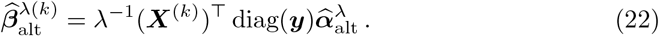

Using (14) and (16), 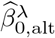 can be computed by combining ***XX***, ***y*** and 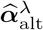. Since, at this stage, these quantities are all available at the CC, the CC is able to compute 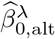.

#### Algorithm 2

RidgeLog-V: Ridge Logistic Regression from Vertically partitioned data with no penalty on the intercept

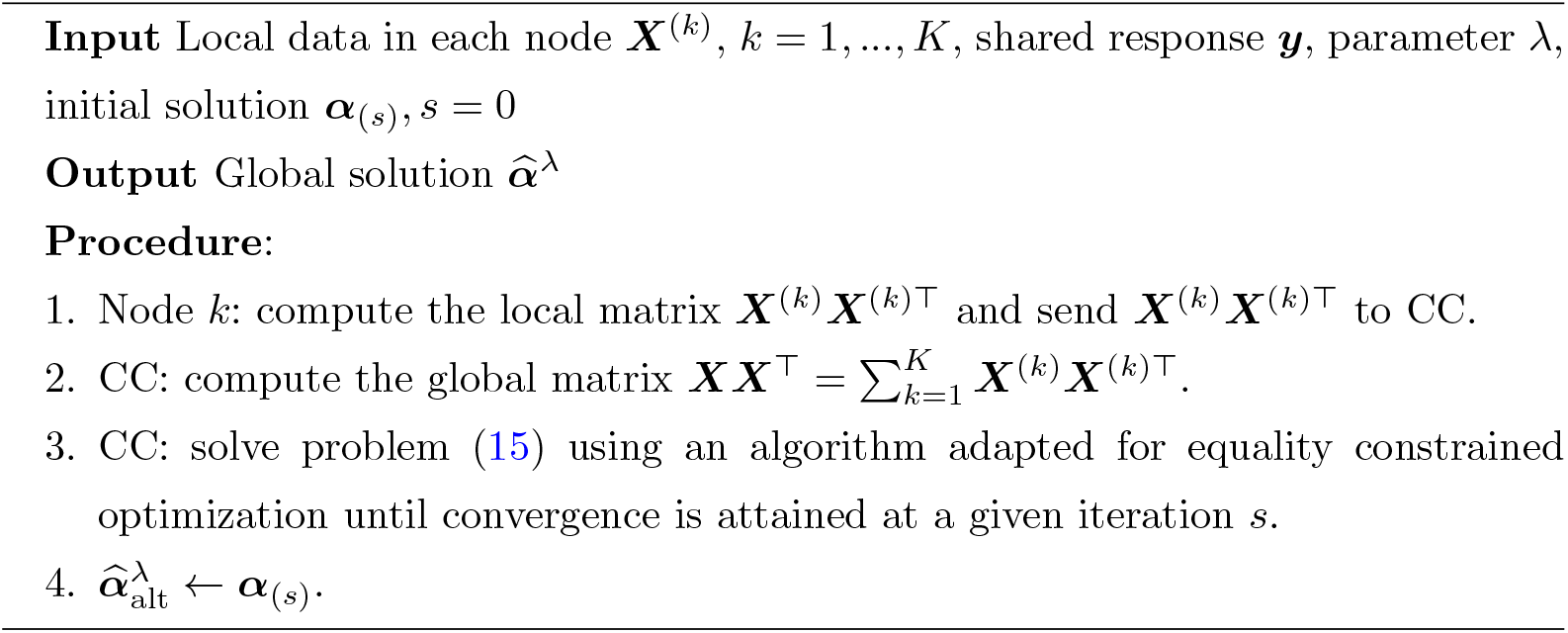

The R implementation of RidgeLog-V (covering objectives (2) and (3)) along with an example comparing its results to those obtained in a centralized setting using the glmnet R package [13] and to those produced by the VERTIGO algorithm, is available at https://github.com/OpenLHS/RevisitingVERTIGO.

## 4 Discussion

To conduct logistic regression analyses in the context of vertically partitioned data, the algorithms VERTIGO and VERTIGO-CI were identified and evaluated with respect to confidentiality, communication efficiency, and comparability to pooled methods.

Unlike in fields where data partitioning may result from limitations in computing resources, the distributed nature of data in many health studies is often due to ethical and legal restrictions that prohibit sharing raw data. In this context, one might mistakenly assume that any method avoiding the direct transmission of raw data is *de facto* compliant with these constraints. However, such constraints rather impose that raw data must remain unknowable to any unauthorized party, regardless of the form in which information is exchanged with the original data source. VERTIGO-CI fails to meet this requirement, as it allows an unauthorized party (the CC) to reconstruct raw data, without the need for intensive computing resources, even though individual data was not directly transmitted (notwithstanding the outocome values). VERTIGO-CI thus serves as another salient example that a distributed algorithm is not inherently privacy-preserving.

While retrieving individual data in VERTIGO-CI required only simple vector-matrix operations involving quantities already available at the CC level, highlighting a flaw in the algorithm’s design, the potential confidentiality breach that arises in all discussed algorithms when a participating node possesses only binary covariates is more subtle. In this case, the binary nature of the features allows for the construction of a system of equations based on the shared quantities. The structure of this system of equations can lead to a unique solution, thereby enabling full reconstruction of the individual data. Our paper identifies conditions under which such recovery is possible. Binary covariates are quite common in this type of analysis. Situations involving a mix of binary and continuous predictors, however, will require users to conduct careful assessments before deploying the method. On the other hand, we did not identify any scenarios in which data nodes with more than three continuous covariates were at risk of reverse-engineering.

This highlights the importance of thoroughly investigating potential data leakage risks when developing methods for distributed analysis. Proving confidentiality is challenging, and often the next best approach is to make as many informed attempts as possible to challenge the potential for reverse-engineering. We found that exploring various forms of representation, such as the matrix form used throughout our approach, can help uncover potential vulnerabilities. Overall, a clear and systematic framework for assessing re-identification risk based on shared quantities remains lacking in the current literature.

What is clear, however, is that the privacy-preserving capabilities of a method cannot be evaluated solely based on the confidentiality properties of individual algorithmic operations. Instead, they must be assessed in light of all quantities that may become available to a potentially adversarial party throughout the entire procedure. For example, the VERTIGO paper claims that when a sufficient number of covariates are present, “the [CC] can obtain the global Gram matrix […] from each party and avoid the disclosure of patient data”, noting that since the Gram matrix is constructed from dot products between patient records, “it is not possible to reverse-engineer the product to obtain the values as long as there are enough covariates from individual databases”. While this may hold under typical data distributions (though it can fail in extreme scenarios, such as when covariates are bounded and all entries lie on boundary values) our findings show that this protection breaks down even under typical conditions when parameter estimates are disclosed and one of the data nodes holds only binary covariates. Such claims may be misleading with respect to the perceived safety of the procedure, especially in the absence of a thorough privacy analysis.

We also identified unnecessary communication steps in VERTIGO and have optimized the algorithm’s communication efficiency. While such changes may appear minor, they can significantly impact the practical applicability of the method to health data. Due to the sensitivity of such data, many institutions require manual review of intermediate results before any further communication with other nodes or the CC. As a result, each additional communication step substantially affects the feasibility of the approach and increases the resources needed to implement it.

This work also underscores the importance of understanding the impact of modeling choices introduced by adapting a method for use in a distributed environment—in this case, those related to penalizing the intercept—to ensure that the resulting estimates remain interpretable in the same way as those typically obtained from similar models in a non-distributed setting. We proposed the alternative RidgeLog-V approach that excludes the intercept from the penalty during estimation, thereby avoiding bias in the average predicted probability of the outcome being one toward one-half.

While the current work has identified important limitations to the context in which the method can be used, it does not mean the method should be entirely discarded. We have not identified confidentiality risks when binary features are included but their associated estimates are withheld. We have also not identified any issue in settings involving enough continuous covariates. In fact, penalized logistic regression is particularly well-suited to scenarios with continuous covariates, especially when these variables are correlated, as the penalty helps mitigate multicollinearity and supports more stable estimation of their relationship with a binary outcome [12]. While the notion of enough covariates was also mentioned in the original VERTIGO paper, no quantification was given. By simple observation of the Gram matrix, cases where a node holds only one or two variables can be problematic, but such issues do not seem to arise with at least three continuous variables. However, in the context of analyses that require privacy guarantees, further investigations should be led before sharing estimates associated with continuous covariates when any of the proposed algorithms are employed.

## 5 Conclusion

We identified confidentiality issues with VERTIGO-CI, concluding that this extension should not be used in settings involving confidential feature data. We also demonstrated the risk of individual data leakage when parameter estimates associated with binary features are shared following the execution of VERTIGO. For use with continuous feature data, we proposed an optimized version of the algorithm to reduce communication costs and introduced an alternative formulation, the RidgeLog-V algorithm, that excludes the intercept from the penalty term. This modification is consistent with standard applications of penalized logistic regression and facilitates a more meaningful interpretation of the intercept.

## Data Availability

All data produced in the present work are contained in the manuscript and available online.

https://github.com/OpenLHS/Distrib_analysis/tree/main/Vertically_distributed_analysis/logistic_regression_penalized

## List of abbreviations

CC: Coordinating center
HDRN: Health Data Research Network
CI: Confidence interval

## Supplementary information

See Additional file 1.

## Declarations

### Ethics approval and consent to participate

Not applicable

### Consent for publication

Not applicable

### Availability of data and materials

The code used in this article is available at https://github.com/OpenLHS/RevisitingVERTIGO.

### Competing interests

The authors declare that they have no competing interests.

### Funding

This work was supported by the Natural Sciences and Engineering Research Council of Canada – Discovery Grant; the Health Data Research Network Canada, an initiative funded by the Canadian Institutes of Health Research; and the Chaire de recherche en informatique de la santé de l’Université de Sherbrooke. Marie-Pier Domingue and Simon Lévesque both received a scholarship from the Natural Sciences and Engineering Research Council of Canada.

### Author contribution

M.P.D., J.F.E., A.B., and F.C.L. conceived and designed the study. M.P.D., J.F.E., and F.C.L. wrote the manuscript. The mathematical derivations were conducted by M.P.D. and F.C.L., with contributions from S.L. and J.P.M. Implementations were carried out by M.P.D., J.F.E., and J.P.M. All authors read and approved the final manuscript.

## Acknowledgments

Not applicable

---

Additional file 1 - *Revisiting VERTIGO and VERTIGO-CI: Identifying confidentiality breaches and introducing a statistically sound, efficient alternative*

## Appendix A Notation

**Table A1.**
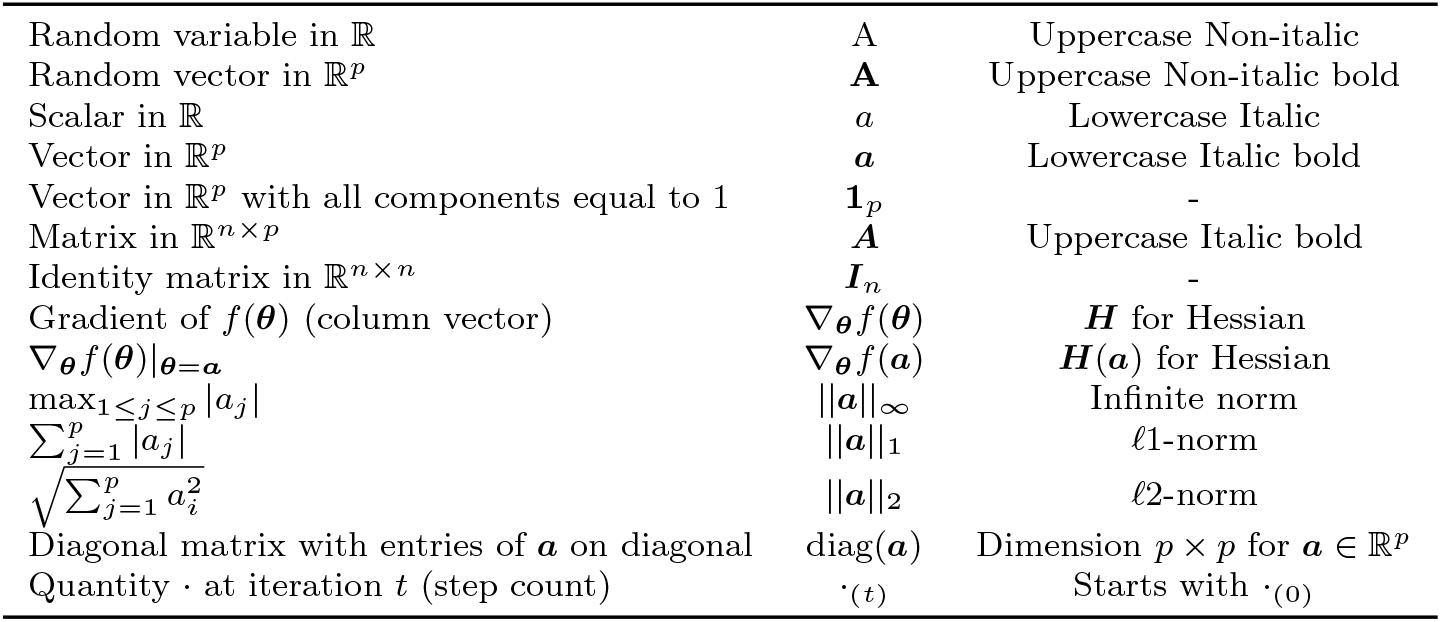
Glossary for general notation conventions.

**Table A2.**
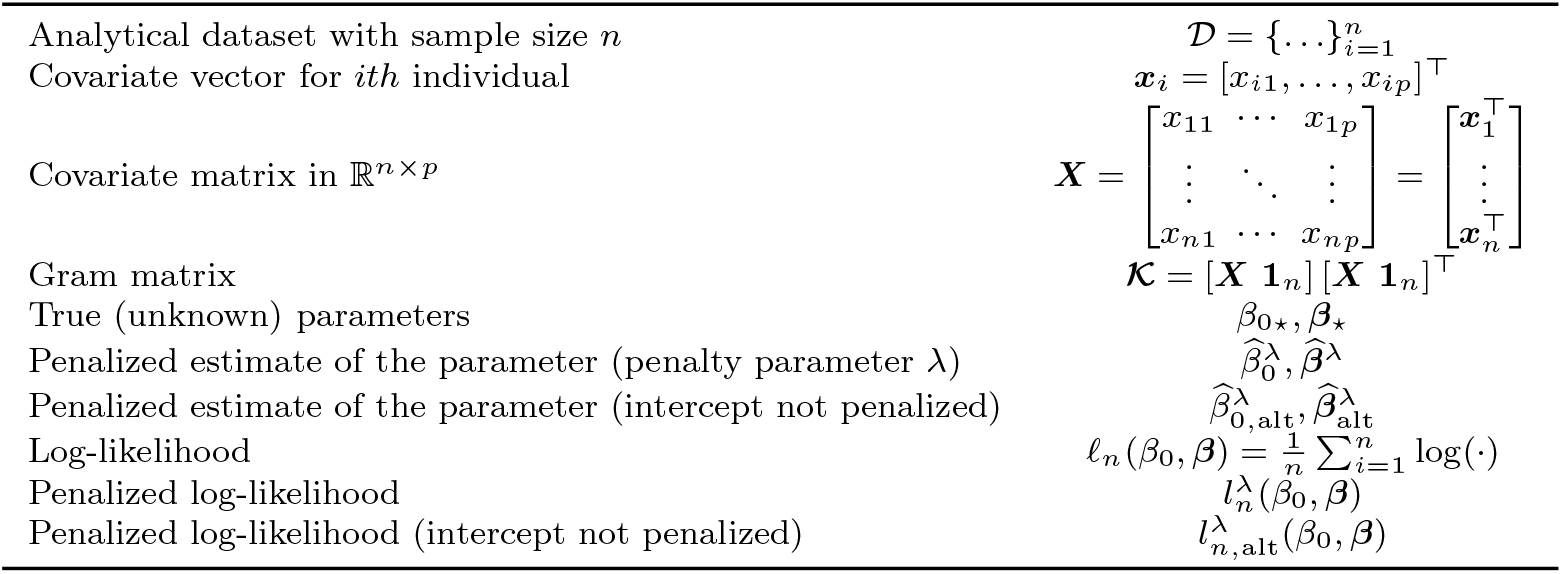
Glossary for quantities that pertain to the regression settings.

**Table A3.** Glossary for quantities specific to the vertical setting.

|  |  |
| --- | --- |
| Number of covariate-nodes | $K$ |
| Number of covariates at covariate-node $k$ | $p^{(k)}$ |
| Covariate matrix at covariate-node $k$ | $\mathbf{X}^{(k)}$ |
| Dual parameter estimates | $\hat{\boldsymbol{\alpha}}^\lambda$ |
| Penalized estimate associated with covariate-node $k$ | $\hat{\boldsymbol{\beta}}^{\lambda(k)}$ |

## Appendix B VERTIGO and VERTIGO-CI Algorithms

Algorithm 3 provides the original VERTIGO algorithm [1], while algorithm 4 provides the original VERTIGO-CI algorithm [2]. Some adjustments have been made to the representation of the original algorithms using the harmonized notation to facilitate the comparison. We note that constrained optimization technique should be considered when numerically applying the Newton-Raphson steps because the components of the parameter ***α*** are constrained in (0, 1).

### Algorithm 3

VERTIGO Original [1]

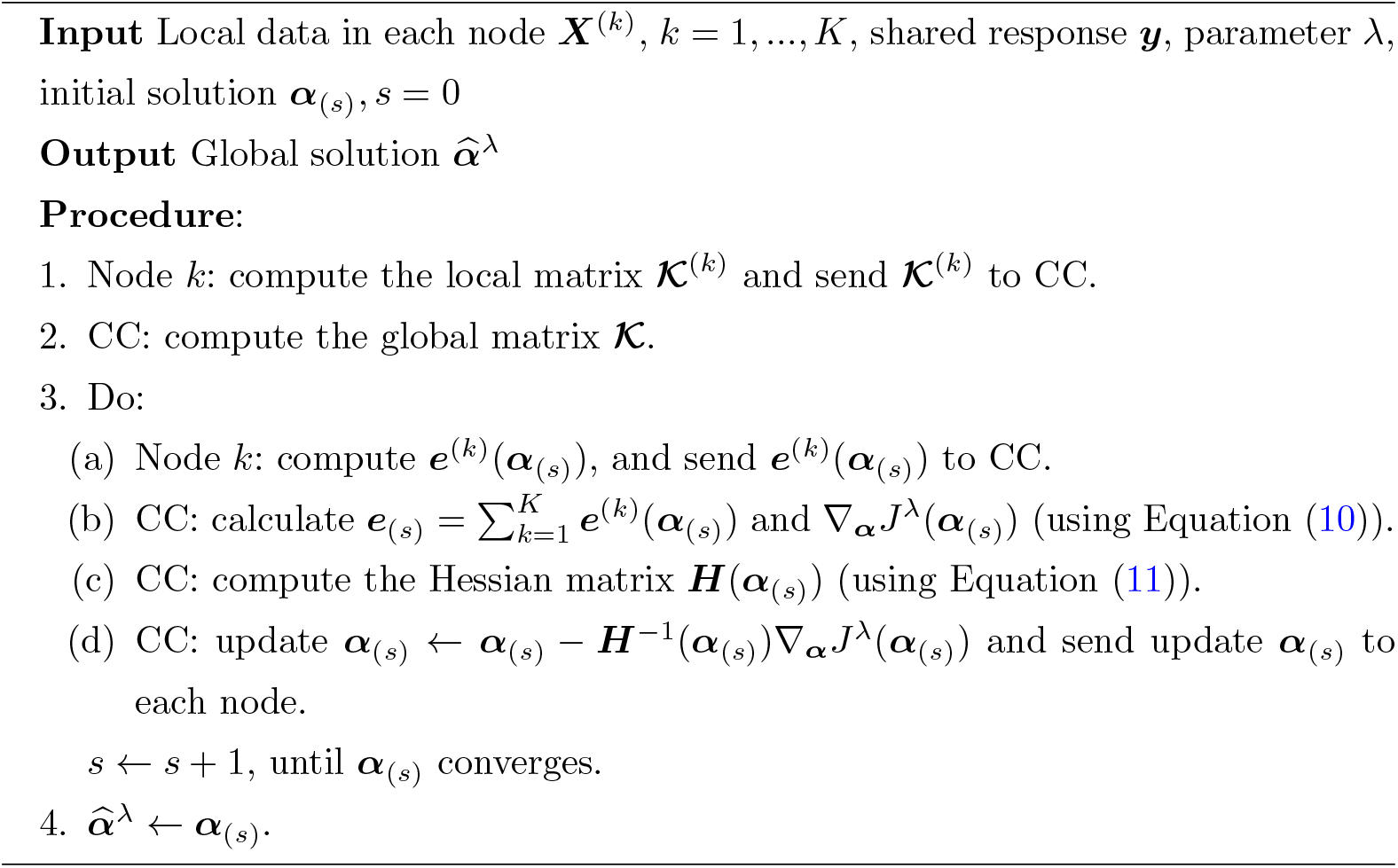

### Algorithm 4

VERTIGO-CI Original [2]

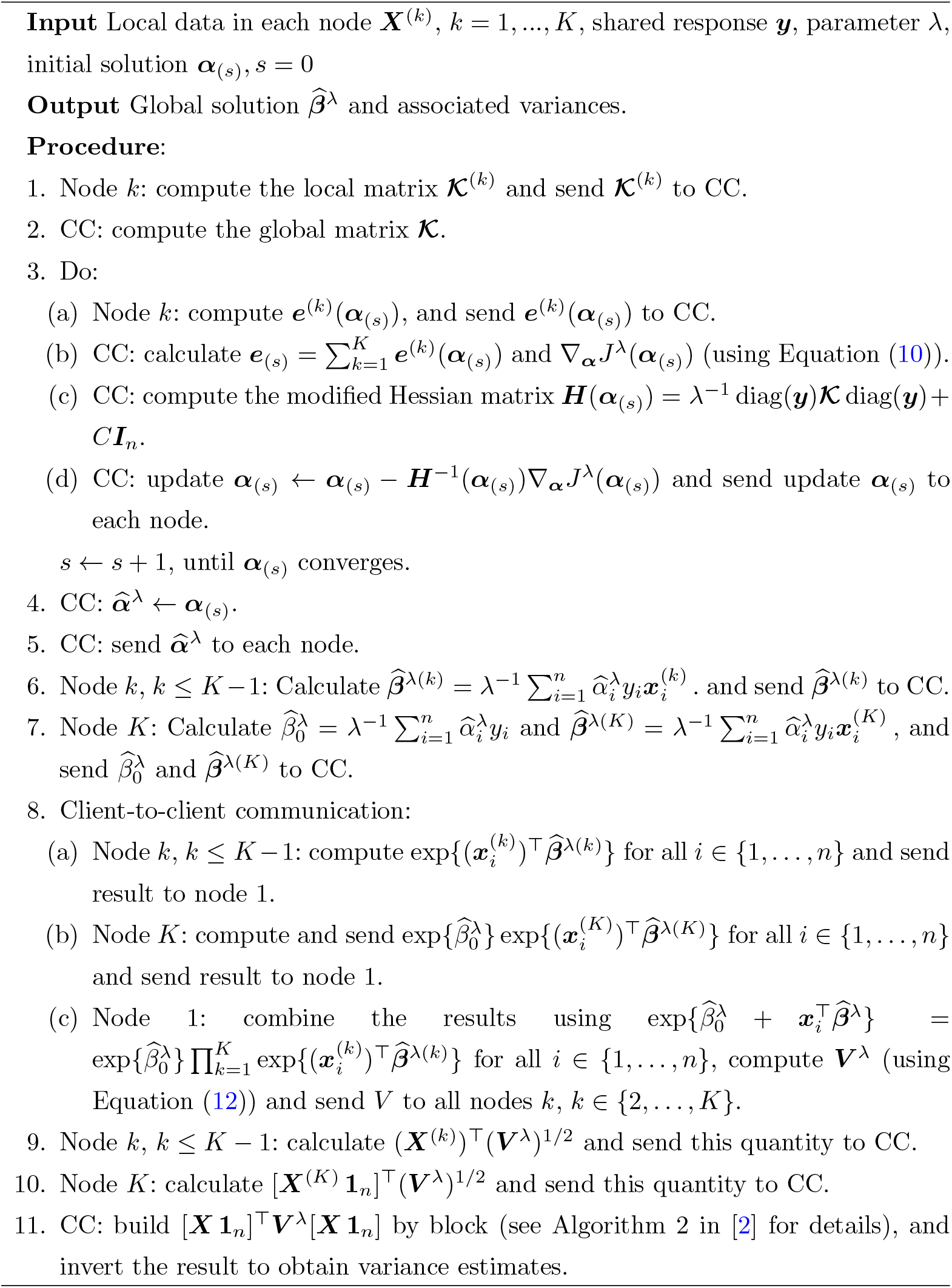

## Appendix C Equivalence between matrix and expanded notation in dual optimization

To obtain the expression of 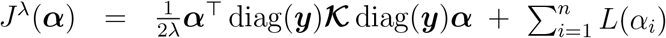, it suffices to note that

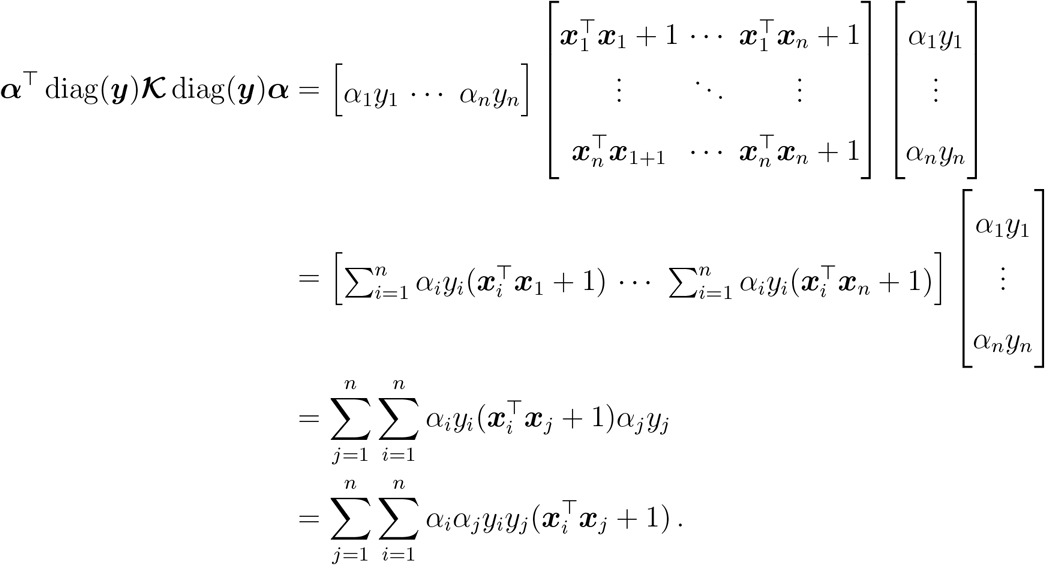

## Appendix D Mathematical development for the dual problem of the ridge logistic regression maximum log-likelihood problem with no penalty over the intercept

### Proposition 1

*Consider any D* = *{*(***x***_1_, *y*_1_), …, (***x***_*n*_, *y*_*n*_)*}, with y*_*i*_ *∈ {−*1, 1*}*, ***x***_*i*_ *∈* ℝ^*p*^ *for i* = 1, …, *n and n ≥* 2. *Assume that* 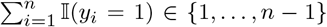 *(i*.*e*., *there exists at least one i such that y*_*i*_ = 1, *and at least one j such that y*_*j*_ = *−*1*). Then, for any λ >* 0, *there exists a unique solution* 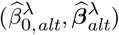 *to the optimization problem*

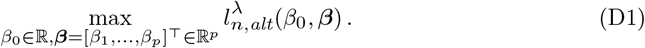

*Furthermore*,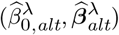*satisfies*

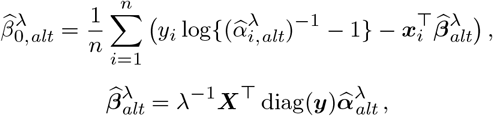

*where* 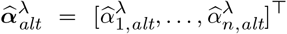 *is the unique solution to the following minimization problem:*

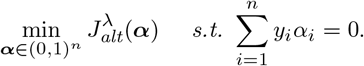

*Proof of Proposition 1*. We begin by showing that the optimization problem in (D1) has a unique finite solution. To do this, we first compute that

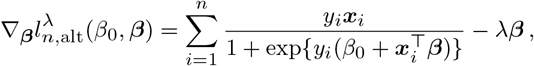

and that

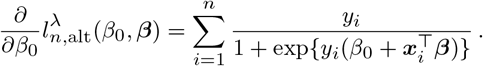

From this, we obtain that

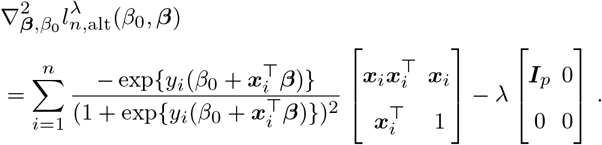

Hence, 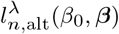 is strictly concave in (*β*_0_, ***β***). Therefore, if 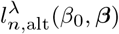 admits a maximizer, it must be unique. To conclude that the optimization problem in (D1) has a unique solution, it remains to show that 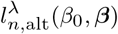 indeed has a maximizer.

To establish this, it suffices to show that 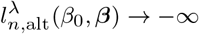 as max{|*β*_0_|, ∥***β***∥_∞_*} → ∞*, where we denote the infinite norm ||***a***||_∞_ := max_1≤*j*≤*p*_ |*a*_*j*_| for *a ∈* ℝ^*p*^. Observe that the loglikelihood term is bounded above by 0. Therefore, if ∥***β***∥_∞_ *→ ∞*, then regardless of whether |*β*_0_| *→ ∞* or |*β*_0_| *< ∞*, the penalty term *λ****β***^⊤^ ***β*** dominates, and we have 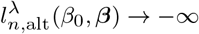.

Now suppose ∥***β***∥∞ ↛ ∞ but max*{*|*β*_0_|, ∥***β***∥_∞_*} → ∞*, which implies that |*β*_0_| *→ ∞* while ∥***β***∥_∞_ remains bounded. Because there is at least one *y*_*i*_ = 1 and at least one *y*_*j*_ = *−*1, there exists at least one term in the log-likelihood of the form log 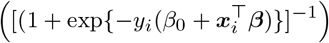 that diverges to *−∞* as |*β*_0_| *→ ∞* (since 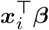remains bounded), while the others remain bounded from above by 0. Therefore, 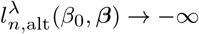 in this case as well.

The conclusion of the above discussion is that for any *λ >* 0, there exists a unique solution 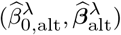) to the optimization problem in (D1). This concludes the proof of the first part of the proposition. The rest of the proof is dedicated to show the second part of the proposition.

Since 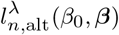 is strictly concave and two times continuously differentiable, 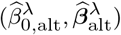 is a stationary point of 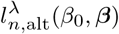, i.e.,

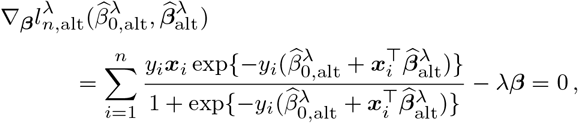

and

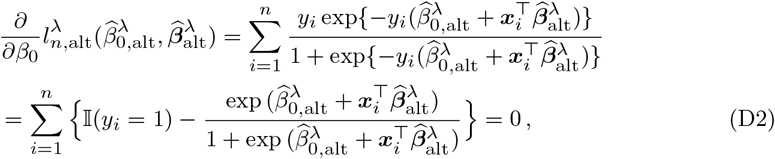

where, on the last line, 𝕀 (*A*) denotes the indicator function taking the value 1 if *A* is true, and 0 otherwise.

Using this result, we will next show that the search space in the optimization problem at (D1) can be narrowed to a compact set (see (D5) below).

From the first equation, we deduce that

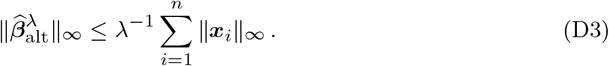

Rearranging the equation for 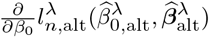 gives

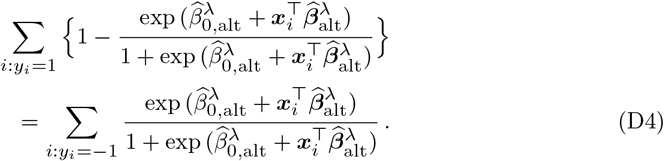

From the latter equation, we will derive in turns a lower bound and an upper bound for 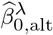. Starting with the upper bound, let *n*_1_ = #*{i* : *y*_*i*_ = 1*}* and *n*_−1_ = #*{i* : *y*_*i*_ = *−*1*}*. Since we have assumed *n*_1_ *≥* 1, we have

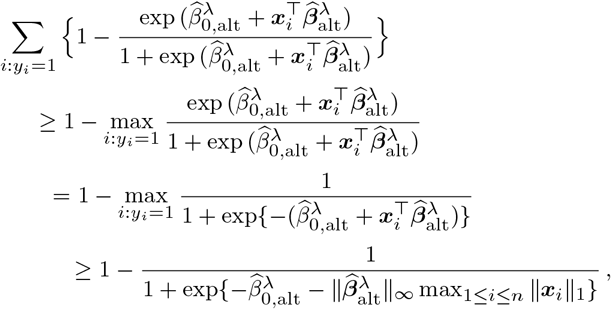

where 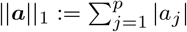 for *a ∈* ℝ. On the other hand, the term on the second line of (D4) satisfies

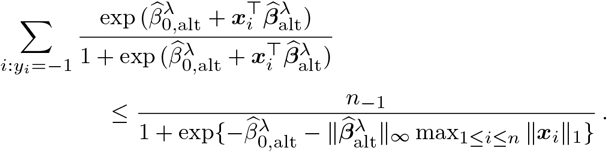

Consequently,

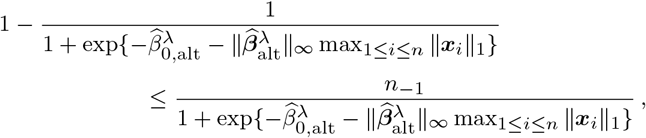

which implies that

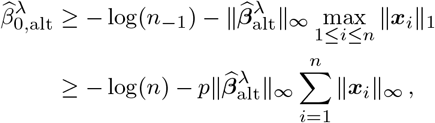

where, to obtain the last line, we used the fact that *n*_−1_ *≤ n*, that for any ***a*** *∈* ℝ^*p*^, ∥***a***∥_1_ *≤ p∥****a***∥_∞_, and that 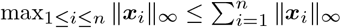.

Since from similar arguments we can show that 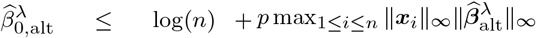, we conclude using (D3) that

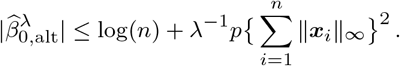

To summarize, we have just shown that the solution 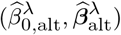 of (D1) lies in the compact set 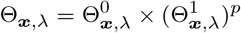, with

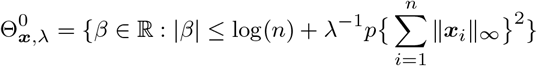

and

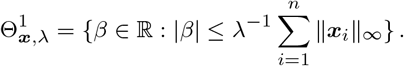

Consequently, the search space ℝ *×* ℝ^*p*^ in the optimization problem at (D1) can be restricted to Θ_***x***,*λ*_, and the optimization problem at (D1) is equivalent to

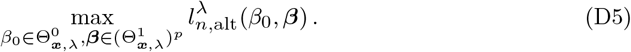

Now from the relationship

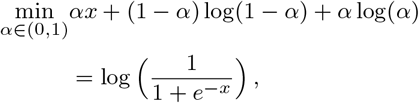

since for any 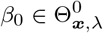 and 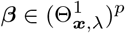, we have

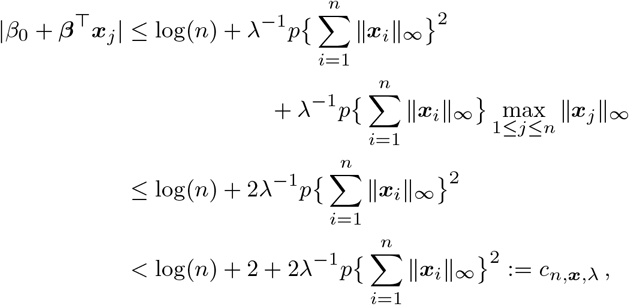

where we used the fact that 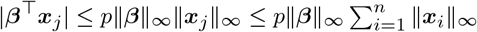, we conclude that

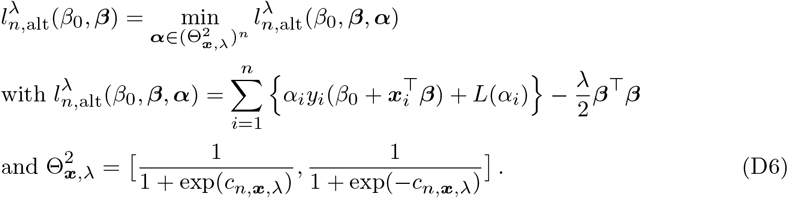

We recall *L*(*α*_*i*_) = (1 *− α*_*i*_) log(1 *− α*_*i*_) + *α*_*i*_ log(*α*_*i*_). Since the solution ***α***(*β*_0_, ***β***) of the optimization problem 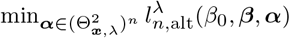 satisfies

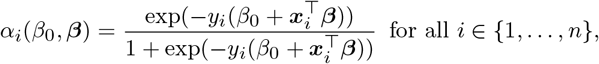

and as the optimal 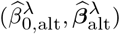 are such that

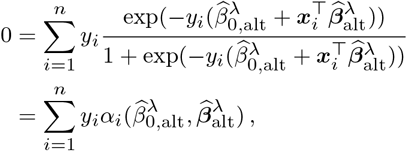

(see (D2)), then, the search space 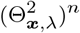 can be further narrowed to

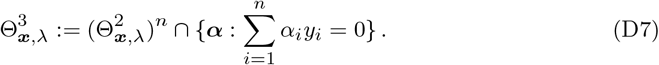

Since for any 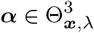 the function 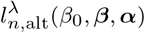 satisfies

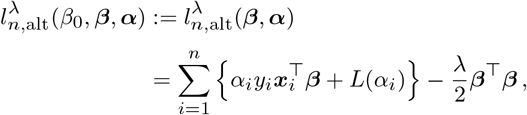

the optimization problem in (D5) can be expressed as

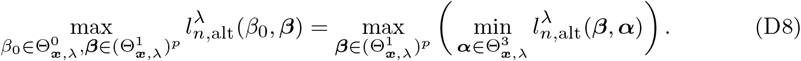

Now for any fixed ***β*** the function 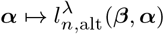 is convex, and for any fixed ***α*** the function 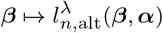 is concave. Since 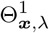 and 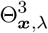 are compact convex sets, we can apply Sion’s minimax theorem [3] and swap the max and the min in the above equation and conclude that

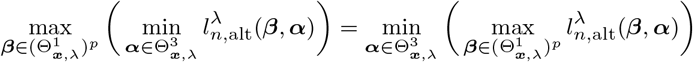

The inner problem can be solved exactly, by equating to 0 the gradient of 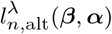 with respect to ***β***. The solution ***β***(***α***) of this equation is given by the following equation:

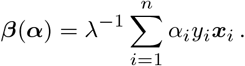

Plugging the expression of ***β***(***α***) into the definition of 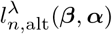 yields 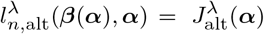,and therefore

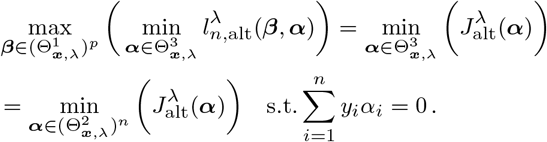

To obtain the last line, we used the definition of 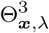 in (D7).

The optimization problem on the last line is strongly convex with a linear equality constraint. It therefore has a unique solution provided the feasible set 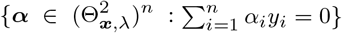 is non empty. This is the case, since, as we have assumed *n*_1_ *≥* 1 and *n*_−1_ *≥* 1 with *n ≥* 2, the point 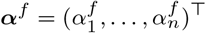 with 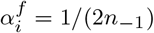 if *y*_*i*_ = *−*1 and 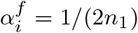 if *y*_*i*_ = 1 is feasible. To see why this is the case, recalling the definition of 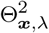 in (D6), it suffices to note that the inequality 1*/*(1 + exp*{x}*) *≤* exp(*−x*) implies that

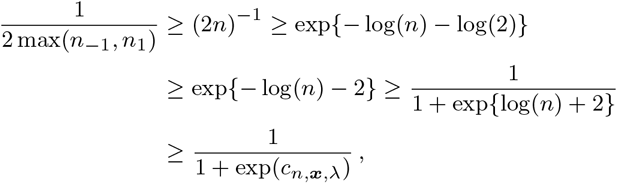

and the inequality 1*/*2 *≤* (1 + exp*{−*2*}*)^−1^ implies that

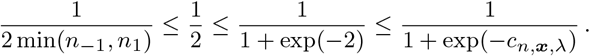

Combining these relationships shows that 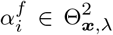 for all 1 *≤ i ≤ n*. The proof of the proposition follows from the fact that, since the solution to the optimization problem

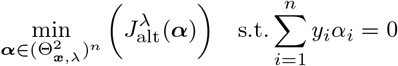

is reached at a stationary point that cancels the Lagrangian, it can be equivalently expressed as

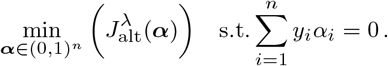

Finally, the expression 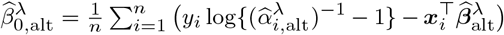 can directly be retrieved using the following:

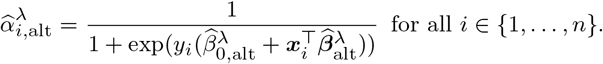

## Appendix E Alternative reverse-engineering procedure

An alternative procedure can be employed to construct the diagonal matrix ***V***^*λ*^ at the CC when executing VERTIGO-CI, which enables the CC to follow the steps outlined in the main text and subsequently reverse-engineer the feature data of all individuals. Recall the definition of the diagonal matrix ***V***^*λ*^, with its entries specified in Equation (12). Also, recall the expression of ∇_***α***_*J*^*λ*^(***α***) (Equation (10)), that 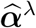 satisfies 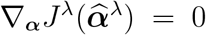, that 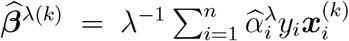 and that 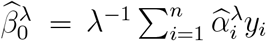.

Using these expression, we derive that the *j*th component of ∇_***α***_*J*^*λ*^(***α***) satisfies

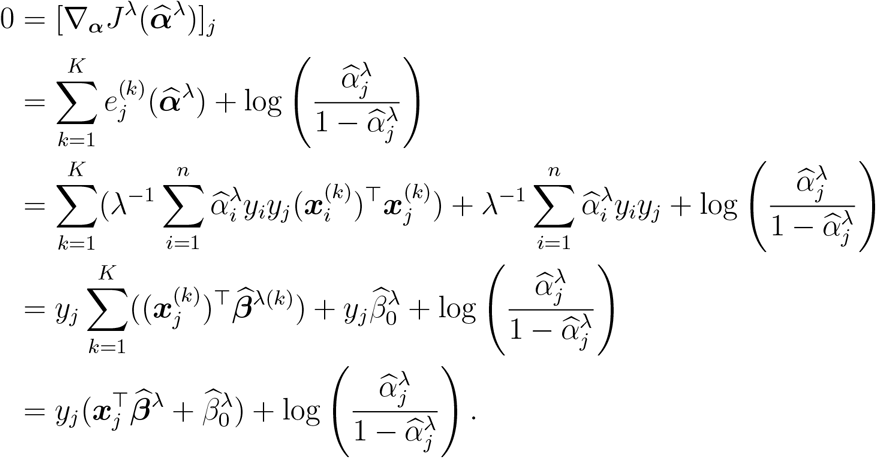

By isolating 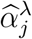, we obtain that

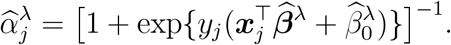

Since 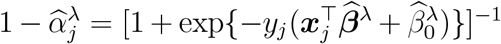, and as *y*_*j*_ ∈ {−1, 1}, we deduce from the latter equation that

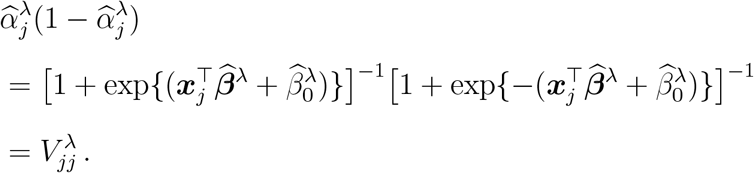

Since 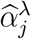 is computed at the CC, the latter can determine each diagonal entry 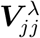 for all *j* ∈ 1, …, *n* and proceed, as outlined in the main text, to reverse-engineer the individual feature data. Notably, this approach allows the CC to reconstruct the matrix ***V***^*λ*^ without requiring access to the response vector ***y***.

## Notes

### Competing Interest Statement

The authors have declared no competing interest.

### Funding Statement

This study was supported by grants from CIHR, NSERC and FRQ(S)

### Summary of Updates

Availability of R code on the github repository.

## References

[1] CanPath.: The Canadian Partnership for Tomorrow’s Health. Available from: https://canpath.ca/en/.

[2] Pfitzner B, Steckhan N, Arnrich B. Federated Learning in a Medical Context: A Systematic Literature Review. ACM Transactions on Internet Technology. 2021;21(2):1–31. 10.1145/3412357.

[3] Brink C, Hansen CR, Field M, Price G, Thwaites D, Sarup N, et al.: Distributed learning optimisation of Cox models can leak patient data: Risks and solutions. Available from: https://arxiv.org/abs/2204.05856.

[4] Agresti A. Foundations of linear and generalized linear models. John Wiley & Sons; 2015.

[5] Slavkovic AB, Nardi Y, Tibbits MM. “Secure” Logistic Regression of Horizontally and Vertically Partitioned Distributed Databases. In: Seventh IEEE International Conference on Data Mining Workshops (ICDMW 2007). Omaha, NE, USA: IEEE; 2007. p. 723–728.

[6] Fienberg SE, Nardi Y, Slavković AB. Valid Statistical Analysis for Logistic Regression with Multiple Sources. In: Protecting Persons While Protecting the People. Berlin, Heidelberg: Springer; 2009. p. 82–94.

[7] Gonçalves C, Bessa RJ, Pinson P. A critical overview of privacy-preserving approaches for collaborative forecasting. International Journal of Forecasting. 2021;37(1):322–342. 10.1016/j.ijforecast.2020.06.003.

[8] Li Y, Jiang X, Wang S, Xiong H, Ohno-Machado L. VERTIcal Grid lOgistic regression (VERTIGO). Journal of the American Medical Informatics Association. 2016;23(3):570–579. 10.1093/jamia/ocv146.

[9] Kim J, Li W, Bath T, Jiang X, Ohno-Machado L. VERTIcal Grid lOgistic regression with Confidence Intervals (VERTIGO-CI). AMIA Summits on Translational Science Proceedings. 2021;p. 355–363.

[10] Moncada-Torres A, Martin F, Sieswerda M, Van Soest J, Geleijnse G. VANTAGE6: an open source priVAcy preserviNg federaTed leArninG infrastructurE for Secure Insight eXchange. AMIA Annual Symposium Proceedings. 2021;2020:870–877.

[11] Kuo TT, Gabriel RA, Koola J, Schooley RT, Ohno-Machado L. Distributed cross-learning for equitable federated models - privacy-preserving prediction on data from five California hospitals. Nature Communications. 2025;16(1). 10.1038/s41467-025-56510-9.

[12] Šinkovec H, Heinze G, Blagus R, Geroldinger A. To tune or not to tune, a case study of ridge logistic regression in small or sparse datasets. BMC Medical Research Methodology. 2021;21(199). 10.1186/s12874-021-01374-y.

[13] Friedman J, Tibshirani R, Hastie T. Regularization Paths for Generalized Linear Models via Coordinate Descent. Journal of Statistical Software. 2010;33(1):1–22. 10.18637/jss.v033.i01.

[14] R Core Team.: R: A Language and Environment for Statistical Computing. Vienna, Austria. Available from: https://www.R-project.org/.

[15] Camirand Lemyre F, Lévesque S, Domingue MP, Herrmann K, Ethier JF. Distributed Statistical Analyses: A Scoping Review and Examples of Operational Frameworks Adapted to Health Analytics. JMIR Medical Informatics. 2024;12(1). 10.2196/53622.

[16] Le Cessie S, Van Houwelingen JC. Ridge Estimators in Logistic Regression. Journal of the Royal Statistical Society Series C (Applied Statistics). 1992;41(1):191–201. 10.2307/2347628.

[17] Fu A, Narasimhan B, Boyd S. CVXR: An R Package for Disciplined Convex Optimization. Journal of Statistical Software. 2020;94(14):1–34. 10.18637/jss.v094.i14.

[18] Boyd SP, Vandenberghe L. Convex optimization. Cambridge, United Kingdom: Cambridge university press; 2004.

## References

[1] Li Y, Jiang X, Wang S, Xiong H, Ohno-Machado L. VERTIcal Grid lOgistic regression (VERTIGO). Journal of the American Medical Informatics Association. 2016;23(3):570–579. 10.1093/jamia/ocv146.

[2] Kim J, Li W, Bath T, Jiang X, Ohno-Machado L. VERTIcal Grid lOgistic regression with Confidence Intervals (VERTIGO-CI). AMIA Summits on Translational Science Proceedings. 2021;p. 355–363.

[3] Sion M. On general minimax theorems. Pacific Journal of Mathematics. 1958;8(1):171–176.

